# Derivation of Pulse Wave Velocity from Arterial Waveforms in Children

**DOI:** 10.1101/2025.11.14.25338673

**Authors:** Ramya Deepthi Billa, John M. Dagle, Mendi Schmelzel, Karen Johnson, Patrick J. McNamara, Aditya V. Badheka, Michael H. Tomasson, Madhuradhar Chegondi, Sarah E. Haskell, Anastasiia Vasileva, Harald M. Stauss, Melissa L. Bates

## Abstract

Aortic stiffness can be measured using aortic pulse wave velocity (PWV), calculated from peripheral arterial waveforms. However, this method has not been validated in children. We sought to determine whether PWV could be feasibly measured by tonometry and accurately derived from the arterial waveforms available in critically ill infants and children. Applanation tonometry was used to measure brachio-femoral or carotid-femoral PWV (b/cfPWV) in hospitalized hemodynamically stable, pre-pubertal children with an indwelling arterial catheter. Peripheral arterial line waveforms were converted into ascending aortic waveforms by a model transfer function. We found that the effective reflecting distance (EfRD) calculation used in adults overestimated b/cfPWV in children. We derived a new EfRD equation that yielded a better agreement between measured and estimated PWV (R² = 0.30, p < 0.05). Estimating PWV from arterial waveforms data eliminates the need for tonometry, allowing larger sample sizes of infants and children to be included in future cardiovascular disease and development studies.

**NEW AND NOTEWORTHY:** Our study provides a method for non-invasively measuring PWV in children using arterial waveforms. We used gold-standard tonometry to measure brachio/carotid-femoral PWV and derived an equation to estimate effective reflecting distance (EfRD) and PWV from the arterial waveforms in a pediatric critical care setting. We determined that PWV can be accurately estimated from arterial waveforms, providing a new tool for studying aortic stiffness in pediatric patients at risk for cardiovascular disease.

## INTRODUCTION

Aortic stiffness is a reliable and independent predictor of cardiovascular morbidity and mortality in adults [1–4]. In addition, increased aortic stiffness may also explain adverse cardiac outcomes in survivors of prematurity [5] and other populations, but methods of estimating pulse wave velocity (PWV), an index of aortic stiffness, in infants and children have not been validated.

Aortic PWV can be measured by arterial applanation tonometry or oscillometric methods with various devices (e.g., Vicorder, Complior, SphygmoCor, Arteriograph), Doppler ultrasound, and Magnetic Resonance Imaging (MRI) [6–9]. PWV measured by arterial applanation tonometry requires pulse transit time between two pulses (carotid-femoral or brachial-femoral), and the measurement of the pulse transit distance between these two sites. Pulse transit time is calculated from the time difference between the ECG R wave (representing ventricular systole) and the foot of the pulse wave at each pulse wave site (t1 and t2). Pulse transit distance is calculated from the distance difference between the sternal notch (landmark for the aortic valve) and the pulse wave measuring sites (d1 and d2). PWV is calculated as pulse transit distance divided by pulse transit time (d2-d1/t2-t1) [10, 11].

An indirect method of deriving PWV from arterial waveforms involves decomposing the arterial waveform into an ascending aortic waveform and estimating the effective reflecting distance (EfRD) [12–14]. The reliability of the model transfer function for converting peripheral arterial waveforms into the ascending aortic waveform has been studied in adults [15, 16]. The ascending aortic waveform is decomposed into forward and backward waveforms by a triangular aortic flow waveform method [17]. Effective reflecting distance (EfRD) is the distance between the aortic pulse wave origin and the reflecting point on the aortic path. The reflecting point is a site on the aorta where the reflecting aortic waves begin to travel back towards the heart, proposed to be located between the aortic-iliac bifurcation and the origin of the femoral artery [18]. PWV from arterial waveforms is derived from dividing the EfRD by the time delay. Effective reflecting distance estimation has been a topic of debate in the literature [19–22], although several published articles propose methods of estimating the reflecting distance using age, weight, height, BMI, heart rate, and blood pressure as variables [12, 23, 24]. The recent method by Pierce et al. estimated EfRD using an equation that includes age and BMI (EfRD in cm= 0.173*age+ 0.661*BMI+ 34.548) in healthy adults to derive PWV from peripheral brachial arterial waveforms and finger pulse waveforms [12]. This method has not been validated in a pediatric population.

It can be challenging to measure PWV by tonometry longitudinally in large cohorts of children across development due to the need for specialized equipment and personnel, particularly when longitudinal studies spanning development are of interest. Most commercially available arterial tonometry devices are not approved for use in infants and young children. Derivation of PWV from arterial waveforms eliminates the need for tonometry, uses commonly available clinical equipment, and can yield a large dataset of longitudinal samples in pediatric patients across developmental years. The purpose of this study was to compare PWV measured by tonometry with estimated PWV to determine whether PWV can be feasibly measured by tonometry and accurately derived from the arterial waveforms available in infants and children.

## METHODS

### Study population

This study was conducted in Neonatal and Pediatric Intensive Care Units (NICU and PICU) at the University of Iowa Stead Family Children’s Hospital. Approval was obtained from the Institutional Review Board at the University of Iowa. Studies were conducted in accordance with the Declaration of Helsinki, and assent and consent were obtained as required. Simulation scenarios were developed to promote this research project and train personnel in the NICU [25]. We included 0–12-year-old children who were clinically stable with existing arterial lines. We excluded clinically unstable patients, including those on extracorporeal membrane oxygenation, patients with brain death upon exam, those with proven or suspected COVID-19, and those with unrepaired aortic arch and aortic valve abnormalities. Clinical stability was determined by the attending physician in the NICU and PICU. Age, weight, height, BMI, vital signs, primary diagnoses, ventilatory support, sedative medications, vasoactive medications, and arterial line site were recorded.

### Arterial waveform collection

Arterial waveforms were collected in the supine position using applanation tonometry, with a commercially available probe (SPT-301, AD Instruments, Colorado Springs, CO). Waveforms were collected at the sternal notch and preferentially at the carotid artery. In cases where the carotid artery was not accessible, waveforms were collected at the brachial artery. Continuous invasive arterial waveforms were recorded from the existing arterial catheter, with the pressure transducer placed at the level of the right atrium. The transducer was zeroed relative to the atmosphere immediately prior to data collection. ECG waveforms were collected from the patient’s existing leads. Under the supervision of a unit nurse, the arterial transducer and ECG leads were connected to a data acquisition device (PowerLab, ADInstruments, Colorado Springs, Colorado) for 5 minutes, and data were recorded at 1000 Hz. The distance between the two tonometry sites was measured with a standard tape measure (d1 and d2). Each LabChart file was uploaded into the open source Hemolab software package. Peripheral arterial waveforms were converted into ascending aortic waveforms by a model transfer function. Ascending aortic waveforms were converted into forward and backward waveforms in Hemolab. Time delays [Δt(f-b/c)] between these forward and backward waveforms were recorded from Hemolab.

### Measurement of PWV from applanation tonometry

Pulse transit time was calculated as the difference between the time delays between the ECG R wave and the foot of the arterial pulse waves at the two measurement sites (carotid or brachial and femoral) (t2-t1). Pulse transit distance was calculated as the site distance from the measurement site and the suprasternal notch, a landmark for the aortic valve (d2-d1). Aortic pulse wave velocity was calculated as PWV=(d2-d1)/(t2-t1).

### Estimation of PWV using a previously published EfRD equation

Estimated EfRD was derived from the method described by Pierce et al [12] (Hemolab, Harald Stauss Scientific) and the previously published equation:

EfRD (in cm) = 0.173*age+0.661*BMI+ 34.548.

True EfRD was calculated as b/cfPWV * ½ Δt(f-b). The estimated EfRD was then used to calculate PWV using the previously described method.

### New EfRD equation and PWV

True EfRD was computed from tonometry measured b/cfPWV and ½ time delay [b/cfPWV * ½ Δt(f-b/c)]. Best subsets logistic regression analysis was done to derive a new EfRD equation using true EfRD as the dependent variable and anthropometric measurements (weight, height, body mass index BMI), pulse transit distances of pulse waves 1 and 2 (d1 & d2), vital signs [heart rate (HR), systolic blood pressure (SBP), diastolic blood pressure (DBP), and mean arterial pressure (MAP)] as independent variables to obtain an new estimated EfRD equation. Results from this equation were used to calculate PWV in Hemolab.

### Statistical analysis

Data are presented as mean±SD, unless otherwise stated. Arterial and tonometry waveforms were reviewed by the study’s principal investigator (MLB) for data quality. True EfRD and tonometry-measured PWV were compared with estimated EfRD and estimated PWV, respectively, using Pearson correlation, intraclass correlation (ICC), and Bland-Altman analyses. Best subsets regression was used to derive a new EfRD equation. The variables that yielded the best R^2^ value and a Mallows’ Cp value that was closest to the number of variables were included in the final equation. Data analysis was done using Minitab (State College, PA), and significance was set *a priori* as p<0.05.

## RESULTS

A total of sixty (60) patients in NICU and PICU with existing arterial lines were included in the study (53% male, median age of 4.2 years, range 0-11.8 years). Four patients had central (umbilical) arterial lines, and 56 patients had peripheral (radial or femoral) arterial lines. The sample included 10 neonates (0-27 days), 15 infants (28 days-12 months), 3 toddlers (13 months-24 months), 11 preschoolers (2-5 years), and 21 school-aged children (6-11 years). Sixty-seven percent (67%) of children were born at term gestational age. Twenty-nine (48%) patients had a prior diagnosis of congenital cardiac disease, and 24 (40%) patients were mechanically ventilated. Patient characteristics are presented in Table 1. Carotid and/or brachio-femoral pulse wave velocity (b/cfPWV) was measured in 55 patients. The final regression analysis included 39 patients with sufficient data quality, free from artifacts.

**Table 1.** Patient Characteristics.

|  | Number of patients (%) |
| --- | --- |
| Sex |  |
| Female | 28 (47) |
| Male | 32 (53) |
| Age* |  |
| Neonates (0 days-27 days) | 10 (17) |
| Infants (28 days-12 months) | 15 (25) |
| Toddlers (13 months-24 months) | 3 (5) |
| Early childhood (2 years-5 years) | 11 (18) |
| Middle childhood (6 years-11 years) | 21 (35) |
| GA at birth |  |
| < 37 weeks | 8 (13) |
| > 37 weeks | 40 (67) |
| Unknown | 12 (20) |
| Respiratory support |  |
| Mechanical ventilation | 24 (40) |
| NIPPV | 4 (7) |
| High flow nasal cannula | 2 (3) |
| Low flow nasal cannula | 8 (13) |
| Room air | 22 (37) |
| Medications |  |
| Vasoactive medications | 15 (25) |
| Sedative medications | 22 (37) |
| Congenital cardiac disease |  |
| Yes | 29 (48) |
| No | 31 (52) |
| *Age stages according to NICHD Pediatric Terminology. NICHD, National Institute of Child Health and Human Development. NIPPV=Non-Invasive Positive Pressure Ventilation. |  |

### Measured PWV compared to the calculated PWV from a previously published EfRD equation

True EfRD derived from PWV*½ Δt(f-b/c) demonstrated poor correlation with estimated EfRD (Figure 1, R^2^=0.03, p=0.16). Bland-Altman analysis showed a mean difference of 34.7 ± 7.6 cm (Figure 2). As a result, measured PWV and calculated PWV were also poorly correlated (R^2^=0.05, p=0.09), and Bland-Altman analysis of measured PWV and calculated PWV revealed a mean difference between the two measurements of 4.9 ± 3.6 m/s and an interclass correlation of −0.67 (−0.81—0.45, 95%CI, p>0.99).

**Figure 1.**
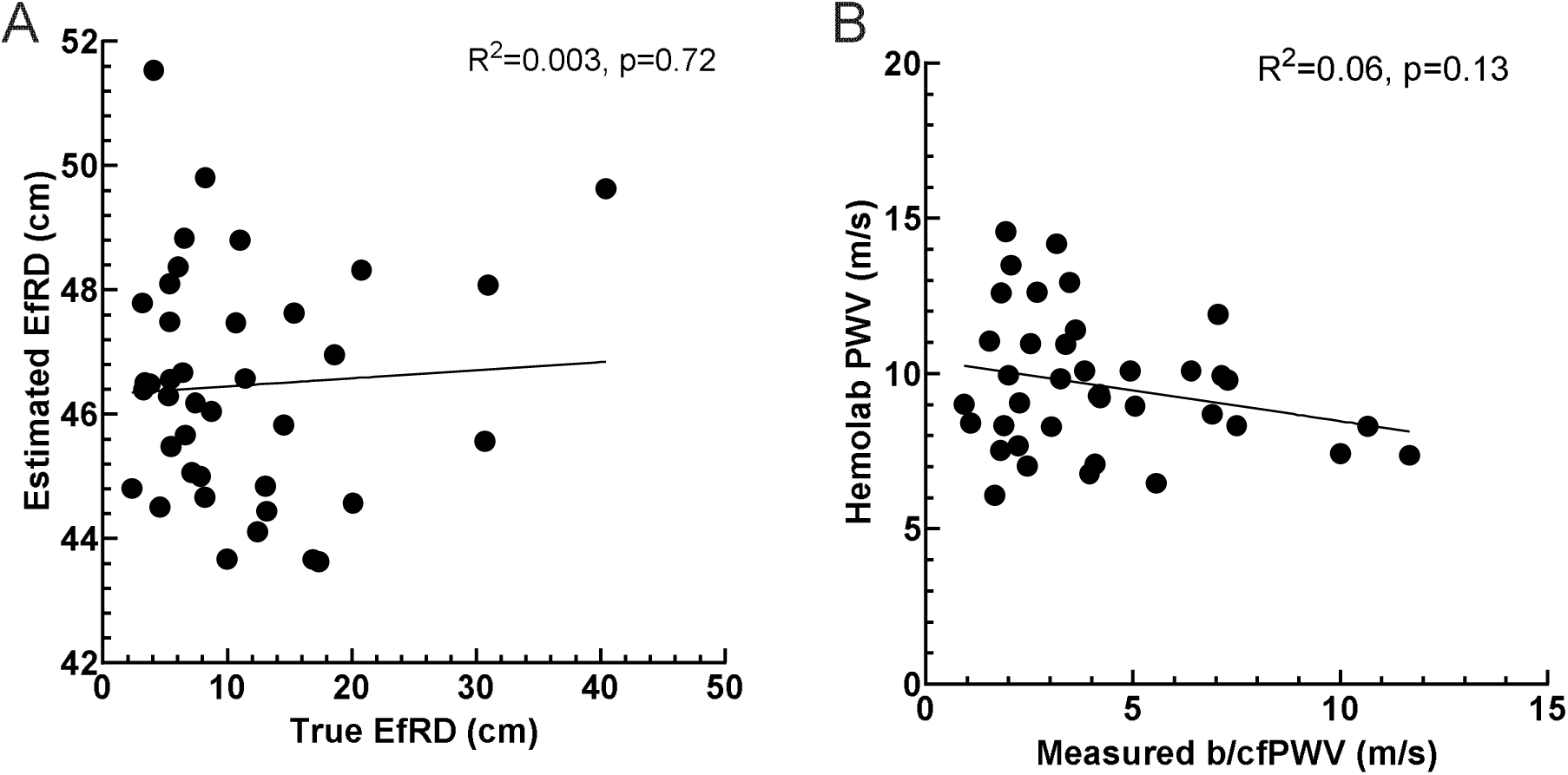
Pearson Correlation. (A) Correlation between true EfRD [derived from the measured PWV*½ Δt(f-b/c)] and estimated EfRD [derived from the formula 0.173*age+0.661*BMI+ 34.548]. (B) Correlation between measured PWV and estimated PWV [derived from the estimated EfRD].

**Figure 2.**
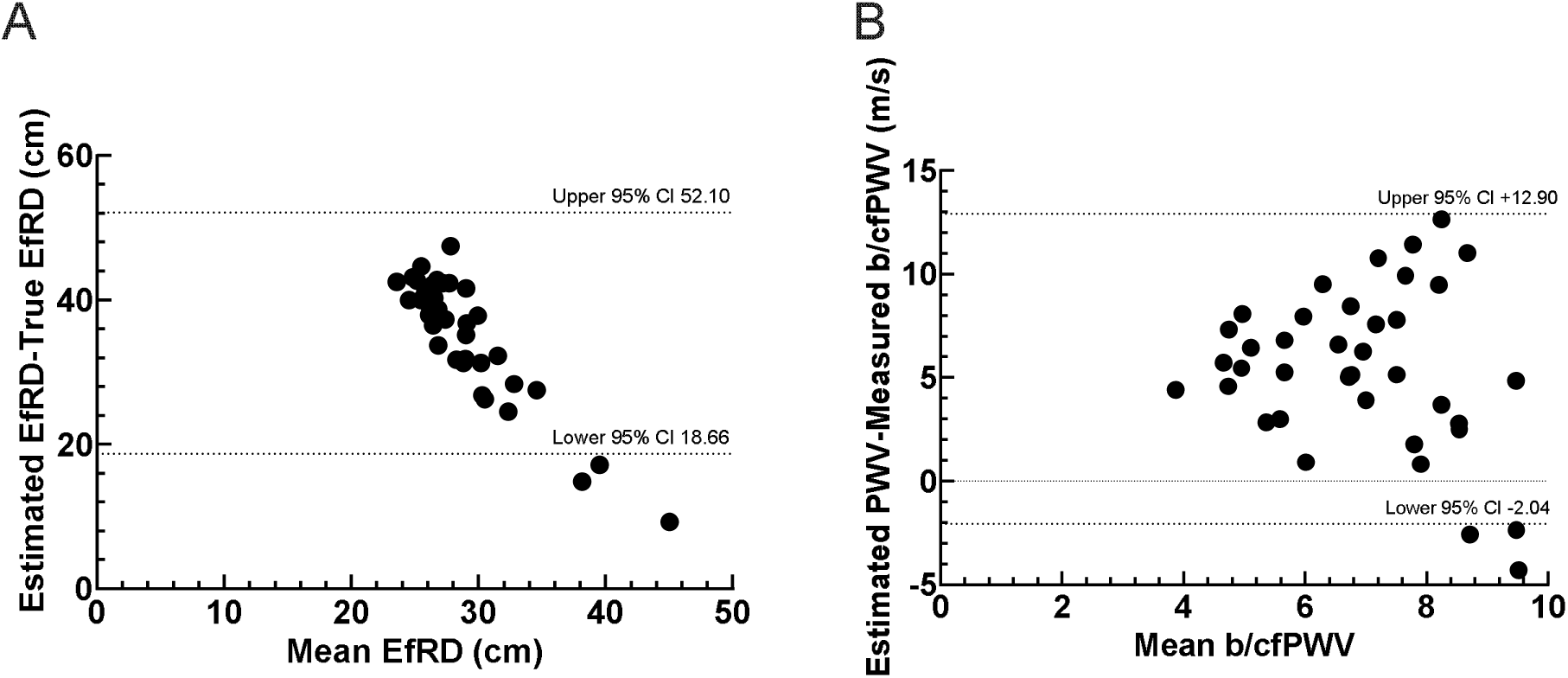
Bland-Altman plots of the differences between estimated EfRD [derived from the equation EfRD (cm) = 0.173*age + 0.661*BMI + 34.548] and true EfRD, and between estimated PWV [derived from the estimated EfRD] and measured PWV from the mean EfRD (A) and mean PWV (B). Dashed lines indicate the 95% confidence interval.

### Measured b/cfPWV compared to calculated PWV from the new EfRD equation

Best subsets regression yielded the following equation for the estimation of EfRD:

EfRD (cm)= (31.1 [if female], 29.9 [if male]) - 0.412*Length (cm) - 0.1448*HR (bpm) + 0.104*SBP (mmHg)+ 1.023*DBP (mmHg)-0.591*MAP (mmHg)+ 0.0926*SBP (percentile) + 0.518*d1 + d2 (cm).

True and estimated EfRD values, measured PWV, and calculated PWV values are shown in Table 2. The correlations between true and estimated EfRD and measured and calculated PWV are shown in Figure 3. There was a strong correlation between true and estimated EfRD (Pearson’s R^2^=0.47, p<0.0001) and a good correlation between measured PWV and calculated PWV (Pearson’s R^2^=0.30, p=0.0002). Bland-Altman plot analysis showed better agreement between the two estimates of EfRD, with a mean difference of 0.08 ± 6.2 cm. The mean difference between the measured and calculated PWV was 0.19 ± 2.3 m/s (Figure 4) and an interclass correlation of 0.53 (0.26-0.73 95% CI, p=0.0002).

**Figure 3.**
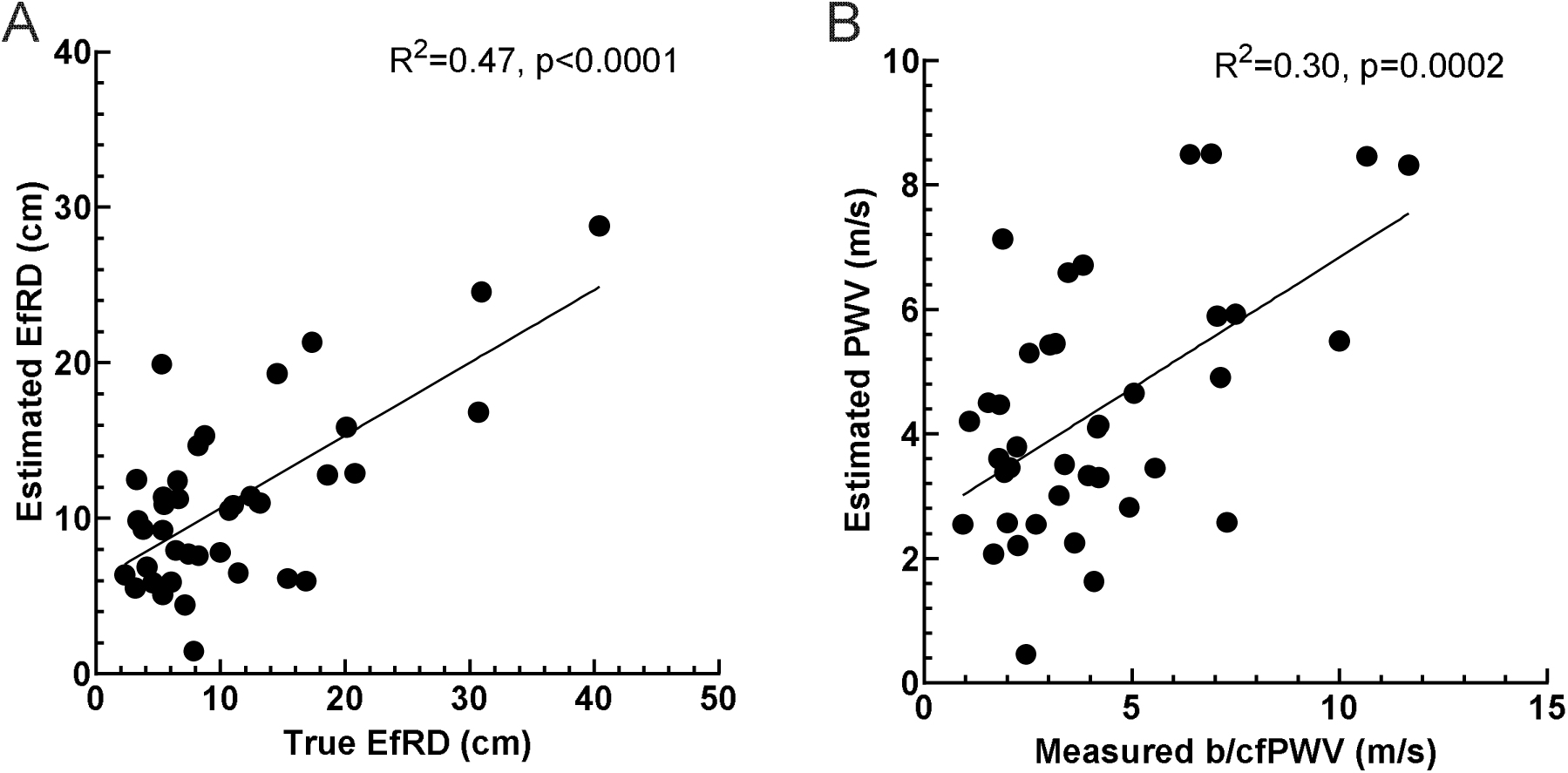
Pearson Correlation. (A) Correlation between true EfRD [derived from the measured b/cfPWV*½ Δt(f-b/c)] and estimated EfRD [derived from the equation EfRD (cm)= 31.1 (if female), 29.9 (if male) - 0.412* Length (cm) - 0.1448*HR (bpm) + 0.104*SBP (mmHg) + 1.023*DBP (mmHg) - 0.591*MAP (mmHg) + 0.0926* SBP percentile + 0.518*d1+d2 (cm)]. (B) Correlation between measured b/cfPWV and estimated PWV [derived from estimated EfRD].

**Figure 4.**
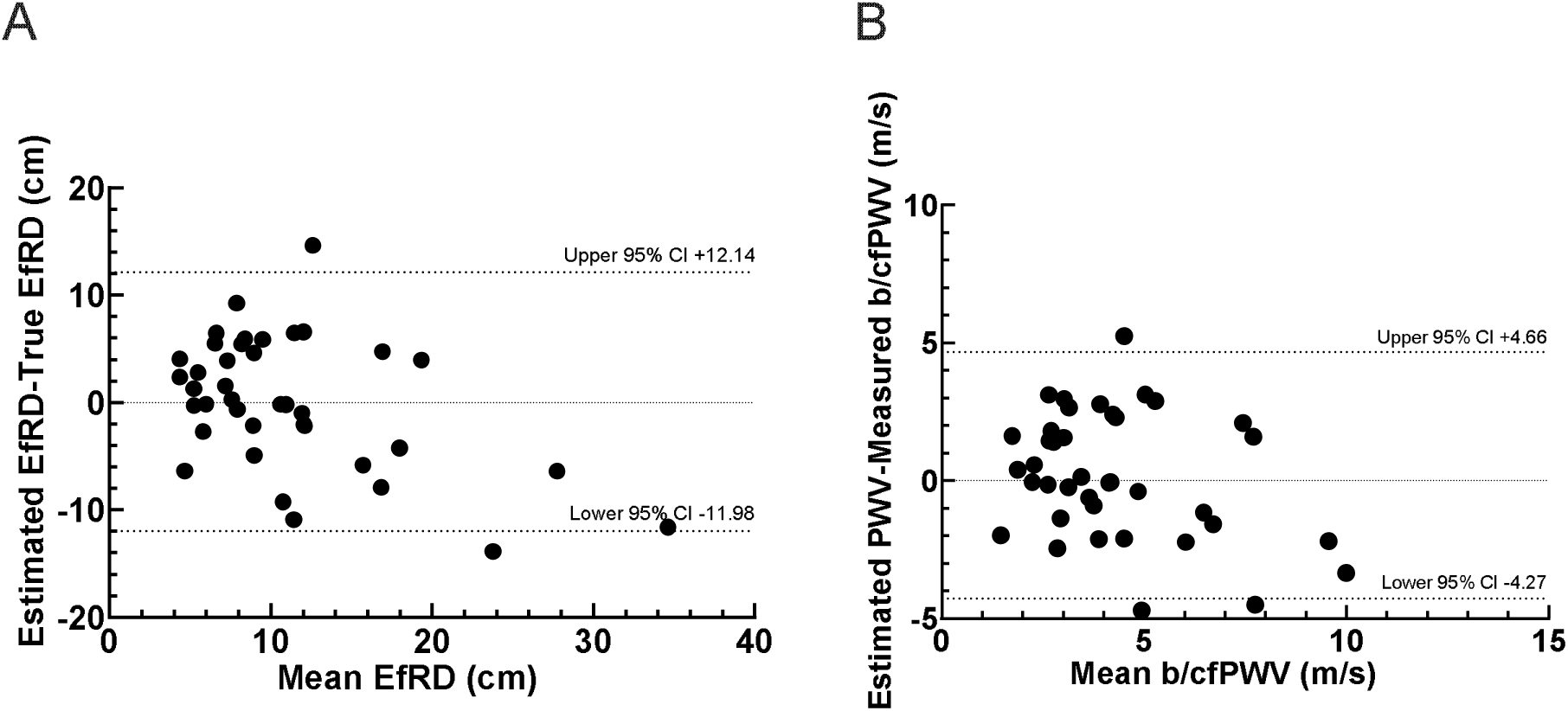
Bland-Altman plots of the differences between estimated EfRD [derived from the equation EfRD (cm)= 31.1 (if female), 29.9 (if male) - 0.412* Length (cm) - 0.1448*HR (bpm) + 0.104*SBP (mmHg) + 1.023*DBP (mmHg) - 0.591*MAP (mmHg) + 0.0926* SBP percentile+ 0.518*d1+d2 (cm)] and true EfRD, and between estimated PWV [derived from the estimated EfRD] and measured b/cfPWV from the mean EfRD (A) and mean PWV (B). Dashed lines indicate the 95% confidence interval.

**Table 2.** Anthropometric and vital signs data.

|  | Mean + SD | Range |
| --- | --- | --- |
| Age (years) | 4.17±4.28 | 0.02-11.81 |
| Weight (kg) | 17.85±17.14 | 3.20-104.60 |
| Height (cm) | 91.59±35.01 | 45.00-160.50 |
| Body mass index (kg/m<sup>2</sup>) | 17.43±4.33 | 11.98-40.61 |
| Systolic BP (mmHg) | 103±19 | 68-145 |
| Diastolic BP (mmHg) | 51±12 | 24-83 |
| Mean arterial pressure (mmHg) | 71±14 | 45-111 |
| Heart rate (beats/minute) | 119±30 | 73-175 |
| Measured bfPWV (m/s) | 4.19±2.66 | 0.93-11.67 |
| Measured cfPWV (m/s) | 4.56±1.27 | 2.85-7.02 |
| Estimated PWV (m/s) | 4.39±2.02 | 0.46-8.50 |
| True EfRD (cm) | 11.08±8.43 | 2.32-40.42 |
| Estimated EfRD (cm) | 11.16±5.77 | 1.47-28.81 |

Kg-kilograms, cm-centimeters, kg/m<sup>2</sup>-kilogram/meter<sup>2</sup>, m/s-meter/second, BP-Blood Pressure, mmHg-millimeter of mercury, b/cfPWV-Brachio/carotid-femoral Pulse Wave Velocity, EfRD-Effective reflecting distance, SD-Standard Deviation.

## DISCUSSION

Here, we describe a method for estimating aortic pulse wave velocity in infants and children and test an existing method to determine its appropriateness for use in children. We found that measured and calculated PWV were not well correlated, due to poor correlation between the estimated and true EfRD. By employing a best-subset regression model, we identified variables that yielded more accurate estimates of EfRD and improved estimates of PWV in infants and children.

Arterial tonometry requires specialized equipment that may not be approved for use in infants and young children, whereas PWV can be estimated using waveforms obtained from standard clinical equipment.

Because arterial tonometry is highly sensitive to motion artifacts, we performed this initial study in the intensive care unit, where patients are most likely to have arterial catheters and receive some degree of sedation to limit movement artifacts. The advantage of using non-invasive arterial waveforms in the future to derive PWV is that data can be collected over a more extended time period, ensuring that only waveforms without motion artifact are included in the analysis. Currently available clinical data acquisition platforms make it highly feasible to record high-resolution waveform data from clinical monitors and apply novel physiological tools to derive endpoints that are not routinely collected or calculated clinically [26].

The applicability of the EfRD equation derived from adult patients for measuring PWV in children is limited. The method described by Pierce et al. uses age and BMI in the adult EfRD equation. This study reemphasized that effective reflecting distance would lengthen with age and reported the new finding that EfRD lengthens with increased BMI [12]. Height is used to derive the EfRD, and it was determined that the effective reflecting distance moves distally with increasing height [23, 24]. The new method for deriving EfRD in our study was independent of BMI or age. Body mass index is not routinely used in pediatrics, and the relationship between age and height can be highly variable, particularly in infancy and during peak velocity growth years [27]. It is, therefore, not surprising that estimations derived from adults would not be a good fit for infants and children.

We derived a new EfRD equation that was dependent on the patient’s length, heart rate, and blood pressure. In particular, heart rate and blood pressure are highly age-dependent, with mean arterial pressure approximately equal to post-menstrual age in newborns [28]. Blood pressure continues to increase throughout development [29] and heart rate falls [30]. Given that PWV is highly dependent on heart rate and blood pressure [31, 32], it is expected that including these variables would enhance the accuracy of estimating EfRD in developing children. Including these variables in the EfRD equation improved its correlation with the true EfRD value, thereby yielding more accurate PWV calculations.

Our results are more consistent with the previous studies in adults that used other anthropometric measurements and vital signs to derive EfRD. Weber et al. used an EfRD equation that included only height (EfRD in cm =[body height/4+ 7.28 cm) [23]. Weir-McCall et al. used age, weight, height, heart rate, and systolic blood pressure in adults to derive EfRD as:

Left Carotid Femoral Path length = 61.03+ 0.81*Age(years) + 132.50*height (m) + 0.59*weight (kg) + 0.21*DBP (mmHg) - 0.14*Heart rate (bpm) + (60.51 if female, 68.48 if male).

EfRD dependence on heart rate was attributed to greater sympathetic activity and associated vasoconstriction in adults. EfRD dependence on blood pressure measured here could also be explained by dependence on vascular tone, which increases with age [24], and we do not know the magnitude by which cardiac output or the existence of intra- or extra-cardiac shunt impacts EfRD.

Before the start of the study, we planned to measure carotid-femoral pulse wave velocity exclusively, but found that this was not feasible in some patients. Children were in a critical care setting and were hemodynamically stable at the time of measurement. However, mechanical ventilation, the presence of a cervical collar, and pacifier swallows created artifacts in the carotid waveform, and neck length was not always sufficient to accommodate the tonometer probe. In those cases, brachial-femoral PWV was measured. Carotid-femoral and branchial-femoral pulse wave velocity are correlated in children >3 years old, and the correlation is device-dependent [33]. This relationship has not been systemically studied in children <3 years old. Future studies may aim to compare the feasibility of using these measurement sites and their correlation, particularly in infants and toddlers.

The outcomes of our study are potentially valuable to researchers and clinicians, as measuring PWV can be very challenging in neonatal and pediatric patients. That said, longitudinal measurement of PWV in pediatrics is important. Young adults born prematurely have elevated aortic stiffness [5], but whether this premature stiffening occurs at or immediately after birth, or is the result of premature aging, is not known. We have also reported an unexpected increase in cardiovascular disease in adults with Down Syndrome, who are at an increased risk of hypotension and cerebrovascular disease [34], but the longitudinal development of the conduit vasculature and PWV have not been well-studied. Deriving PWV from the arterial waveform would enable measurements in a large number of patients, longitudinally and across the lifespan, thereby allowing these types of questions to be answered.

## LIMITATIONS

We had patients across all prepubertal age groups, although not enough patients in each group were enrolled to quantify the impact of development on estimated PWV definitively. Although cfPWV is considered the gold-standard measure of arterial stiffness, bfPWV was sometimes used in this study. This methodological difference may limit the comparability of our results with studies that use exclusively cfPWV. We measured PWV in the critical care setting because of the regular use of invasive arterial pressure monitoring and the potential to perform tonometry without movement artifact. However, several factors may affect the absolute PWV value, including cardiac output, shunt status, hypertension, discomfort, mechanical ventilation, sedatives, vasoactive medications, and antihypertensive medications. This study only aimed to associate measured and estimated endpoints, and the values reported here should not necessarily be interpreted as normal for conscious, healthy children. Still, the strong correlation between measured and estimated endpoints suggests the potential for use of a non-invasive method in other pediatric populations, including the ability to relate PWV to clinical factors and outcomes across large patient populations.

Patients had peripheral arterial catheters placed at different sites, including radial and femoral arteries. We also had three patients with umbilical arterial lines. The waveforms from umbilical arterial lines were not converted into ascending aortic waveforms. Instead, they were directly used as central aortic waveforms. Peripheral arterial waveforms were converted into ascending aortic waveforms by a model transfer function. The model transfer function is insensitive to changes in wave transmission delay along the tube, and peripheral capacitance*peripheral resistance [35]. We used the default values in Hemolab for the wave transmission time delay for all arterial lines. The method by Pierce et al. mentioned that the differences in wave transmission time delays are minimal when using brachial versus radial arterial waveforms [12].

## CONCLUSION

We show that PWV can be accurately estimated from arterial waveforms in infants and children, and that calculated velocities correlate well with measured velocities. We report a modified equation for the estimation of EfRD in infants and children. We demonstrate in a cohort of infants and children in the intensive care unit that estimated pulse wave velocity is well-correlated with pulse wave velocity derived from a peripheral arterial waveform. Brachio-femoral PWV was the most feasible method of measuring PWV in critically ill infants and children. Our newly derived EfRD equation, which incorporates body length, blood pressure, and heart rate, improves the accuracy of estimating PWV and can be used to study aortic stiffness in a pediatric population without the need for tonometry. This method will facilitate the study of aortic pulse wave velocity throughout childhood development and can be used to evaluate factors associated with increased aortic stiffness across different clinical settings, including among survivors of prematurity and patients with Down syndrome.

## Data Availability

All data produced in the present study are available upon reasonable request to the authors.

## Author Contribution

All authors participated in the design of the experiments, data collection and analysis, preparation of the manuscript, and have approved the final manuscript for submission

## Disclosures

Dr. Bates is the Founder and CEO of LSF Medical Solutions and Dr. Tomasson serves as Chief Medical Officer. Harald Stauss is the creator of Hemolab. LSF Medical Solutions LLC and Hemolab neither provided nor received funding for the conduct of this work.

## Grants or Funding

None to disclose

## Notes

### Funding Statement

This study did not receive any funding.

### Author Declarations

IRB of the University of Iowa gave ethical approval for this work.

